# A cost-effectiveness analysis of increased quadruple therapy use in heart failure with reduced ejection fraction in Singapore

**DOI:** 10.64898/2026.02.10.26346043

**Authors:** Sameera Senanayake, Audry Shan Yin Lee, Sanjeewa Kularatna, Thin Mar Win, Annie Lee, Yee How Lau, Derek J Hausenloy, Khung-Keong Yeo, Mark Yan-Yee Chan, Raymond Ching Chiew Wong, Seet Yoong Loh, Kheng Leng David Sim, Chow Weien, Kelvin Bryan Tan, Tan Ngiap Chuan, Nicholas Graves

**Author notes:** ***Corresponding author:*** Sameera Senanayake (PhD, MD, MSc, MBBS), Health Services Research & Population Health, Duke-NUS Medical School, Singapore, |.

## Abstract

**Background:** Quadruple therapy, comprising an angiotensin receptor–neprilysin inhibitor (ARNI), β-blocker, mineralocorticoid receptor antagonist (MRA), and sodium–glucose cotransporter 2 inhibitor (SGLT2i), is guideline-recommended for heart failure with reduced ejection fraction (HFrEF). However, uptake in Singapore remains low. This study evaluated the cost-effectiveness of scaling up quadruple therapy from the current 30% uptake to realistic (80%) and stretch (100%) targets.

**Methods:** We developed a decision-analytic model combining a decision tree and Markov structure to simulate clinical and economic outcomes over a 10-year horizon from the Singapore healthcare system perspective. Transition probabilities were estimated using local real-world data for current regimens, and published literature for quadruple therapy. Costs were derived from hospital billing data and drug utilisation patterns. A probabilistic sensitivity analysis (1,000 simulations) assessed uncertainty. The willingness-to-pay (WTP) threshold was S$45,000 per quality-adjusted life year (QALY) gained.

**Results:** Both scale-up scenarios were cost-effective. Compared to current practice, the 80% uptake scenario resulted in an incremental cost of S$2.57M and 110 additional QALYs (ICER: S$23,392/QALY) for 1000 patients over 10 years, while the 100% uptake scenario yielded 137 QALYs at an incremental cost of S$2.88M (ICER: S$21,117/QALY). Under conservative assumptions, both scenarios remained cost-effective. The probability of being cost-effective was 92% (80% uptake) and 96% (100% uptake).

**Interpretation:** Scaling up quadruple therapy for HFrEF in Singapore is highly cost-effective. Implementation strategies to close the treatment gap should be prioritised to improve outcomes and maximise value in heart failure care.

## Introduction

Heart failure with reduced ejection fraction (HFrEF) remains a major cause of morbidity and mortality worldwide, with rising prevalence driven by population ageing and improved survival from cardiovascular disease^1,2^. In Singapore, heart failure accounts for substantial healthcare utilisation, including frequent hospital readmissions and long-term medical management costs, placing an increasing burden on the public health system^3,4^.

Over the past decade, advances in pharmacological therapy have improved survival and quality of life for patients with HFrEF. Guideline-directed medical therapy (GDMT) now recommends the early initiation of quadruple therapy, comprising preferably an angiotensin receptor–neprilysin inhibitor (ARNI), otherwise an angiotensin-converting enzyme inhibitor (ACEi)/angiotensin receptor blocker (ARB), a beta-blocker, a mineralocorticoid receptor antagonist (MRA), and a sodium–glucose cotransporter 2 inhibitor (SGLT2i). Evidence from recent randomised controlled trials, such as PARADIGM-HF^5^, DAPA-HF^6^, and EMPEROR-Reduced^6^ suggests combined use of these agents reduces all-cause mortality and heart-failure-related hospitalisations compared with conventional therapy. Implementing quadruple GDMT at optimal doses has been estimated to reduce 12-month mortality by approximately 10% in absolute terms^7^. Global modelling further suggests that full implementation of quadruple therapy could prevent up to 1.2 million deaths annually, with the greatest impact projected in regions such as Southeast Asia, the Eastern Mediterranean, Africa, and the Western Pacific^7^.

Despite strong clinical evidence and clear guideline recommendations, real-world uptake of quadruple therapy remains suboptimal both globally and in Singapore. Our previous research identified that only 30% of an eligible patient cohort in Singapore (excluding patients with guideline-listed absolute contraindications), were receiving all four drug classes as of the year 2022. Possible barriers to optimal implementation may include physician and patient inertia, knowledge gaps, cost concerns and healthcare system gaps in service provisions for early initiation and up-titration such as limited availability of clinic appointments.

Given the increasing emphasis on efficiency and cost-effectiveness in Singapore’s healthcare system^8^, it is important to assess whether increasing adherence to quadruple therapy will increase efficiency. While clinical benefits are well-established, the higher drug costs associated with comprehensive GDMT mean we are unsure whether increasing uptake is a cost-effective policy goal.

This study aimed to evaluate the cost-effectiveness of increasing adherence to quadruple therapy for patients with HFrEF in Singapore, from the healthcare system perspective.

## Methods

We developed a decision-analytic model combining a decision tree and a Markov model (Figure 1) using TreeAge Pro 2025 to evaluate the cost-effectiveness of increasing adherence to quadruple therapy for HFrEF in Singapore. The analysis compared current practice with two additional quadruple therapy scenarios. Quadruple therapy was defined as treatment with all four key GDMT drug classes: an ACEi/ ARB/ ARNi; a beta-blocker; an MRA; and a SGLT2i. Based on our previous work, only 30% of an eligible patient cohort in Singapore were prescribed quadruple therapy in 2022. Eligibility for GDMT was determined according to the listed absolute contraindications in the 2021 European Society of Cardiology (ESC) Guidelines for the diagnosis and treatment of acute and chronic heart failure^9^. Patients without contraindications for each drug class were classified as eligible for the corresponding drug class, and those without contraindications for any of the four classes were considered eligible for quadruple therapy. Two improved uptake scenarios were modelled: a possible target of 80% and a stretch target of 100% quadruple therapy adoption among eligible patients.

**Figure 1.**
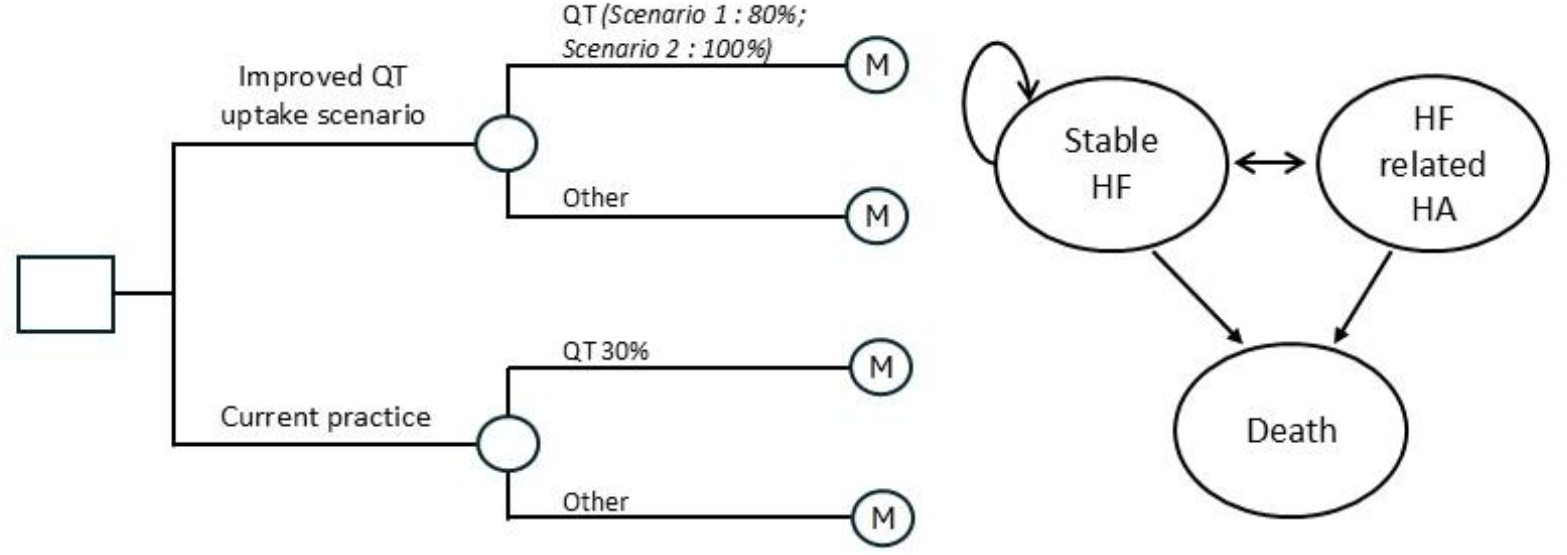
Markov model with the health states and possible transitions

The study was conducted in accordance with a predefined economic evaluation plan and we report our findings following the Consolidated Health Economic Evaluation Reporting Standards (CHEERS) guidelines^14^.

### Target population

The target population comprised patients diagnosed with HFrEF following their first (first as in de novo or incident episode) heart-failure-related hospital admission, with a median age of 70 years. Patients were classified as having HFrEF based on baseline echocardiographic findings, defined as a left ventricular ejection fraction (LVEF) ≤ 40%.

### Model structure

The model was adapted from previously published cost-effectiveness analyses of guideline-directed medical therapy in heart failure^10,11^ and refined through consultation with heart-failure specialists in Singapore. The model comprised two components: a decision tree and a Markov model (Figure 1). The decision tree compared three alternatives: current practice is 30% of eligible patients receiving quadruple therapy, and two competing scenarios reflect realistic (80%) and stretch (100%) uptake targets. Each branch of the decision tree, such as the ‘quadruple therapy’ branch and the ‘other drug regimen’ branch, fed into one of two Markov models. The quadruple therapy branch linked to a model representing the long-term disease trajectory of patients receiving quadruple therapy, while the ‘other’ branch represented patients treated with other drug regimens. The Markov model included three mutually exclusive health states: stable heart failure, heart-failure-related hospitalisation, and death. All patients entered the model in the stable heart-failure state following diagnosis. During each model cycle, patients could remain stable, transition to heart-failure-related hospitalisation, or progress to death. Those who experienced a hospitalisation could either return to the stable heart-failure state or transition to death.

The complete model was simulated over a 10-year time horizon, transitioning in 1-year cycles through the three health states. Effectiveness was evaluated in terms of Quality Adjusted Life Years (QALYs).

*QT-Quadruple therapy; HF-Heart Failure; HA-Hospital admission*

### Data sources

#### Transition probabilities

Separate transition probabilities were estimated for the two cohorts receiving quadruple therapy and ‘other drug regimens’.

For the cohort on ‘other drug regimens’, transition probabilities were derived from the Singapore Cardiovascular Longitudinal Outcomes Database (SingCLOUD). Established in 2014, SingCLOUD is a comprehensive health data platform that integrates information from multiple national sources, including public hospitals and the Ministry of Health^12^. Data quality and integrity are ensured through multiple validation and quality-assurance processes. The other drug regimens cohort included patients receiving combinations such as ACEi/ARB/ARNI + beta-blocker + MRA (21 %), ACEi/ARB/ARNI + beta-blocker + SGLT2i (15 %), ACEi/ARB/ARNI + beta-blocker (21 %), beta-blocker alone (21 %), and no drugs (6 %).

From this dataset, three transition probabilities were estimated for patients who are on ‘other drug regimens’:

1. the probability of heart-failure-related hospitalisation,
2. the probability of all-cause death while in the stable-heart-failure state, and
3. the probability of all-cause death while hospitalised.

Only patients who remained on a consistent drug regimen throughout the follow-up period were included in the analysis to ensure internal validity; those with regimen changes were excluded.

As expected from disease progression patterns, these were modelled as time-dependent transition probabilities. Weibull regression was used to estimate these probabilities, as the Weibull distribution provided a visually good parametric fit to the observed non-parametric survival functions^13^. The Weibull model allows the baseline hazard to increase or decrease over time, offering flexibility in representing dynamic risk^14^. Each Weibull regression model was adjusted for covariates, including age, sex, race, and the presence of comorbidities such as diabetes mellitus, liver disease, cerebrovascular disease, hypercholesterolaemia, chronic lung disease, coronary artery disease, body-mass index > 25 kg/m^2^, diastolic blood pressure > 90 mmHg, systolic blood pressure > 140 mmHg, and chronic kidney disease stage. The resulting λ (scale/rate parameter) and γ (shape parameter) estimates were used to calculate the time-dependent transition probabilities for each outcome (Table 1).

**Table 1.**
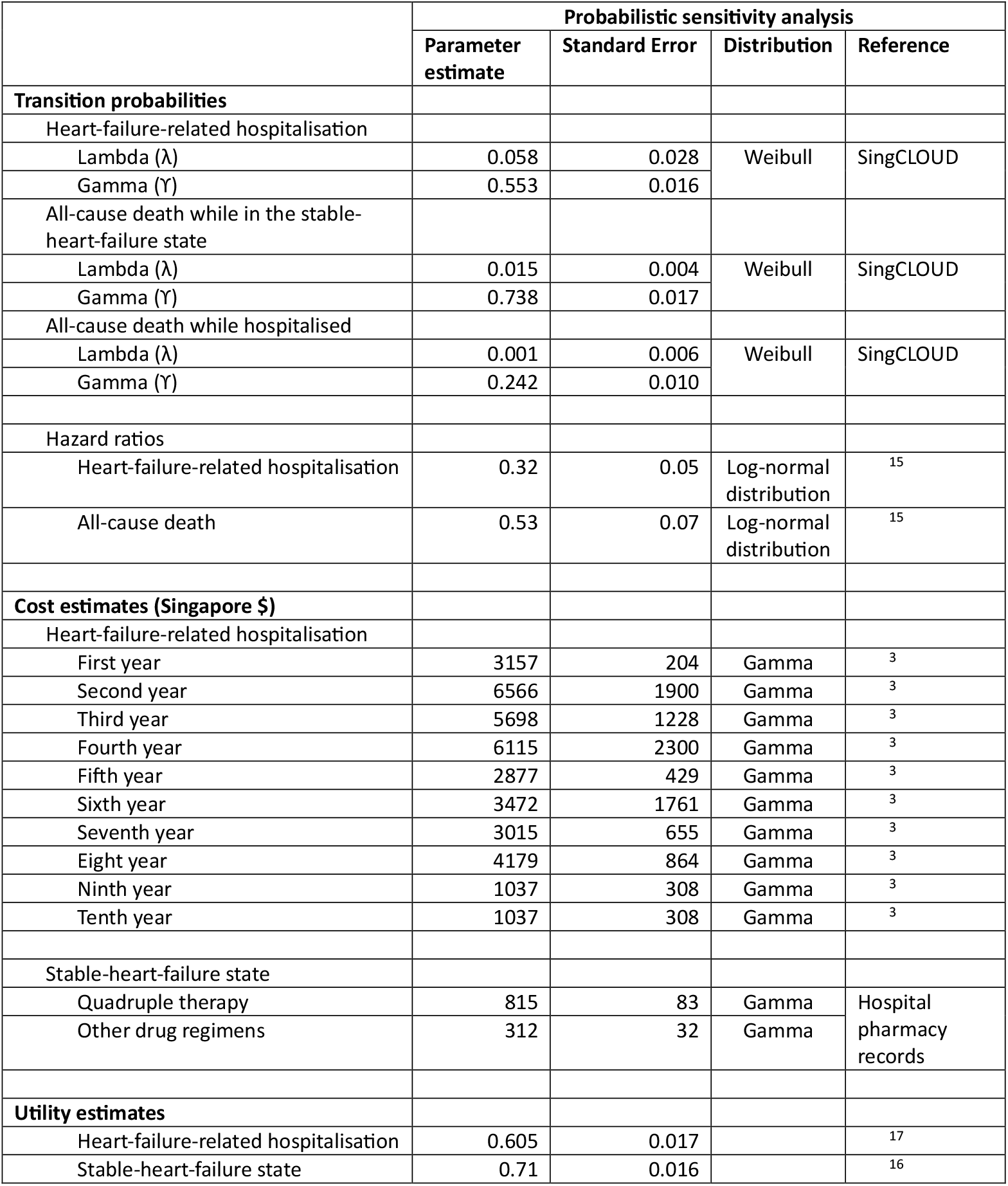
Parameters used in the Markov model.

Due to limitations in patient numbers and follow-up duration, SingCLOUD data were not used to estimate transition probabilities for the cohort receiving quadruple therapy. The use of quadruple therapy increased only after public subsidies were introduced for SGLT2 inhibitors, with 91, 168, and 332 patients on quadruple therapy in 2020, 2021, and 2022, respectively. As SingCLOUD data were available only up to 2022, the small cohort size and limited follow-up period precluded reliable estimation of transition probabilities for this group.

Consequently, transition probabilities for the quadruple-therapy cohort were informed by published evidence. Vaduganathan et al. (2020) conducted a cross-trial analysis that compared comprehensive disease-modifying pharmacological therapy (ARNI, beta-blocker, MRA, and SGLT2 inhibitor) with conventional therapy (ACEi or ARB plus beta-blocker) in patients with chronic HFrEF^15^. The analysis integrated data from three pivotal randomised controlled trials—EMPHASIS-HF (n = 2,737), PARADIGM-HF (n = 8,399), and DAPA-HF (n = 4,744). According to this study, the hazard ratio for heart-failure-related hospitalisation with quadruple therapy versus conventional therapy was 0.32 (95 % CI 0.24–0.43), and the hazard ratio for all-cause mortality was 0.53 (95 % CI 0.40–0.70). These hazard ratios were applied to the λ (rate) parameters estimated for the ‘other-drug-regimen’ cohort, while assuming that the γ (shape) parameter remained constant across treatment groups, as the treatment effect was modelled to proportionally reduce the baseline hazard without altering its time dependency. Because the control arm in the Vaduganathan analysis represented ACEi or ARB plus beta-blocker therapy—whereas the ‘other-drug-regimen’ cohort included a range of regimens (e.g. ACEi/ARB/ARNI + beta-blocker + MRA, ACEi/ARB/ARNI + beta-blocker + SGLT2i, beta-blocker alone, and no GDMT)—we modelled an additional conservative scenario in which the hazard ratios were increased by 50% to account for potential differences in treatment efficacy. This adjustment was not intended to reflect a clinically plausible effect size, but rather to test the robustness of the model under a deliberately pessimistic assumption.

#### Cost estimates

The cost of heart-failure-related hospital admissions was derived from our previous work, which used data from SingCLOUD^3^. This retrospective, longitudinal cohort study followed 1,631 patients with a LVEF<40% for nine years. The annual costs following the first hospital admission were used as model inputs for the cost of heart-failure-related hospitalisation (Table 1).

Our earlier research also identified that patients with HFrEF in Singapore are prescribed heterogeneous pharmacological regimens (e.g. among beta-blocker, the two main agents used are bisoprolol and carvedilol) and are frequently maintained on lower than guideline-recommended dosages. The annual cost of managing stable heart failure was therefore estimated by accounting for both the distribution of prescribed drugs and the average dosage levels observed in clinical practice. For instance, within the ACEi/ARB/ARNI group, approximately 40% of patients were prescribed sacubitril/valsartan (Entresto) and 22% were prescribed valsartan, while the mean daily Entresto dose was approximately 100mg, below the recommended 400mg/day. All drug prices were obtained from hospital pharmacy records, and detailed cost components are presented in Supplementary Table 1. Based on these estimates, the annual cost of managing stable heart failure was $875 for patients receiving quadruple therapy and $138 for those on ‘other drug regimens’.

Additionally, we assumed that achieving the realistic target of 80% quadruple therapy uptake would require an implementation intervention. A one-time cost of $1,000 per patient was incorporated to account for this intervention.

All costs were reported in 2025 Singapore Dollars.

#### Utility estimates

There were no local health utility data so regional and international estimates were used to assign utility weights to each health state. The mean utility values were 0.71 (SD 0.016) for the stable heart-failure state^16^ and 0.605 (SD 0.017) for the heart-failure-related hospitalisation state^17^.

### Model evaluation

In the decision-analytic model, cohorts of patients with HFrEF were simulated through a series of health states over a 10-year time horizon. The cost-effectiveness analysis compared the total accumulated costs and QALYs between current practice and the two quadruple-therapy scale-up scenarios. Both costs and QALYs were discounted at an annual rate of 5%. The analysis was conducted from the perspective of the Singapore healthcare system.

A probabilistic sensitivity analysis (PSA) showed the impact of parameter uncertainty on model outcomes. Probability distributions were assigned to model inputs to capture their underlying variability: beta distributions were applied to transition probabilities and utility values bounded between 0 and 1, while gamma distributions were used for cost parameters to account for the right-skewed nature of healthcare costs. A total of 1,000 Monte Carlo simulations were conducted, with input values randomly sampled from the assigned distributions, see Table 1. For each simulation, the incremental cost-effectiveness ratio (ICER) was calculated as the difference in mean costs divided by the difference in mean QALYs between the scaled-up scenario and current-practice arms. The net monetary benefit (NMB), representing the monetary value of health gains minus the additional costs of the intervention, was estimated using the following equation:

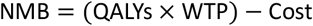

A willingness-to-pay (WTP) threshold of $45,000 per QALY gained was applied^18^. Both average NMB and incremental NMB were calculated, with incremental NMB derived by subtracting the NMB of current practice from that of each scale-up scenario. The scenario yielding the highest NMB was considered the most cost-effective option, as selecting a lower-value alternative would imply an opportunity cost. Final estimates of total cost, effectiveness, ICER, and NMB were derived from the mean outcomes across all PSA simulations, ensuring robustness under parameter uncertainty.

## Results

Table 2 presents the main scenario cost-effectiveness results for the two quadruple-therapy scale-up scenarios over a 10-year time horizon for 1,000 patients. To visualise the joint distribution of cost and effectiveness outcomes, an incremental cost-effectiveness scatterplot was generated (Figure 2), with each point representing the incremental cost and incremental QALYs from one PSA iteration. Compared with current practice, both the realistic (80%) and stretch (100%) uptake scenarios resulted in higher total costs but greater QALY gains. The ICERs were S$23,392 per QALY gained for the 80% uptake scenario and S$21,117 per QALY gained for the 100% uptake scenario with probabilities of cost-effectiveness of 92% (Figure 2 [A]) and 96% (Figure 2 [B]), respectively.

**Table 2.** Cost-effectiveness of the two quadruple-therapy scale-up scenarios, based on 1,000 Monte Carlo simulations.

| | Cost (S\$) | Incremental Cost (S\$) | QALY | Incremental QALY | ICER (S\$/QALY) | Probability of being CE |
| --- | --- | --- | --- | --- | --- | --- |
| Current practice | 6,492,609 |  | 6,771 |  |  |  |
| Scaled up scenarios |  |  |  |  |  |  |
| Quadruple-therapy 80% | 9,058,990 | 2,566,382 | 6,881 | 110 | 23,392 | 92% |
| Quadruple-therapy 100% | 9,375,758 | 2,883,149 | 6,907 | 137 | 21,117 | 96% |

*Results are for 1000 patients over 10 years*

**Figure 2.**
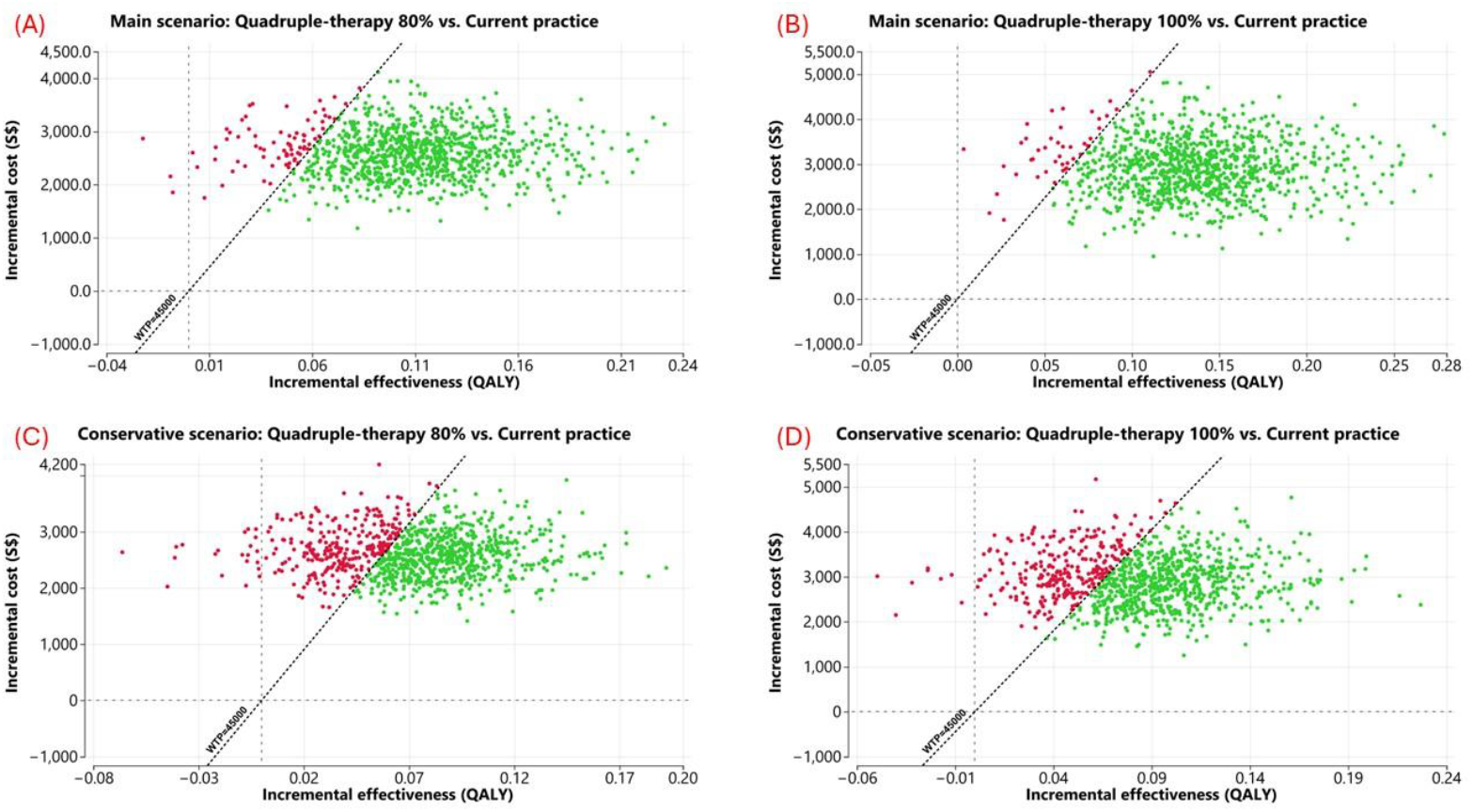
Incremental cost-effectiveness scatterplots for the realistic (80%) and stretch (100%) quadruple-therapy scale-up scenarios, showing results from 1,000 Monte Carlo simulations

Table 3 presents the cost-effectiveness outcomes under a conservative assumption, in which the hazard ratios for quadruple therapy were increased by 50%. Even under these conservative conditions, both quadruple-therapy scale-up scenarios remained cost-effective. The ICERs were S$35,404 per QALY gained for the 80% uptake scenario and S$34,350 per QALY gained for the 100% uptake scenario. The corresponding probabilities of cost-effectiveness were 69% (Figure 2 [C]) and 70% (Figure 2 [D]), respectively.

**Table 3.**
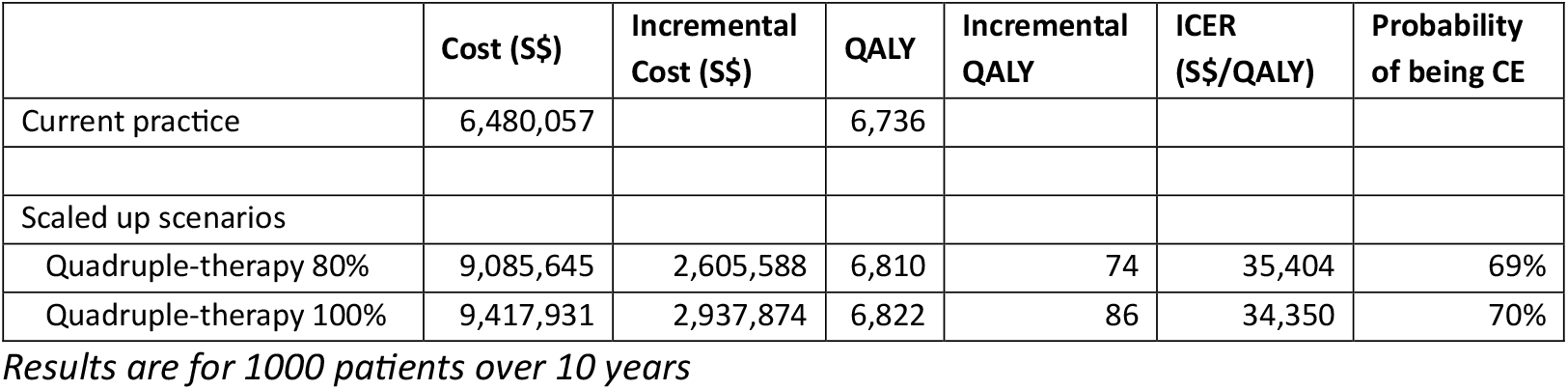
Cost-effectiveness of quadruple-therapy scale-up scenarios under the conservative assumption (hazard ratios increased by 50%), based on 1,000 Monte Carlo simulations Monte Carlo simulations.

Figure 3 presents the incremental cost, incremental effectiveness, and incremental NMB scatterplots for the realistic (80%) and stretch (100%) quadruple-therapy scale-up scenarios under both the main scenario and conservative assumptions in the PSA. Across both analyses, approximately 99% of simulations indicated that the scale-up scenarios were both more costly and more effective than current practice. A net positive NMB was observed in about 97% of simulations under the main scenario and in around 69% of simulations under the conservative scenario, reinforcing that quadruple-therapy scale-up remained cost-effective even under less favourable assumptions.

**Figure 3.**
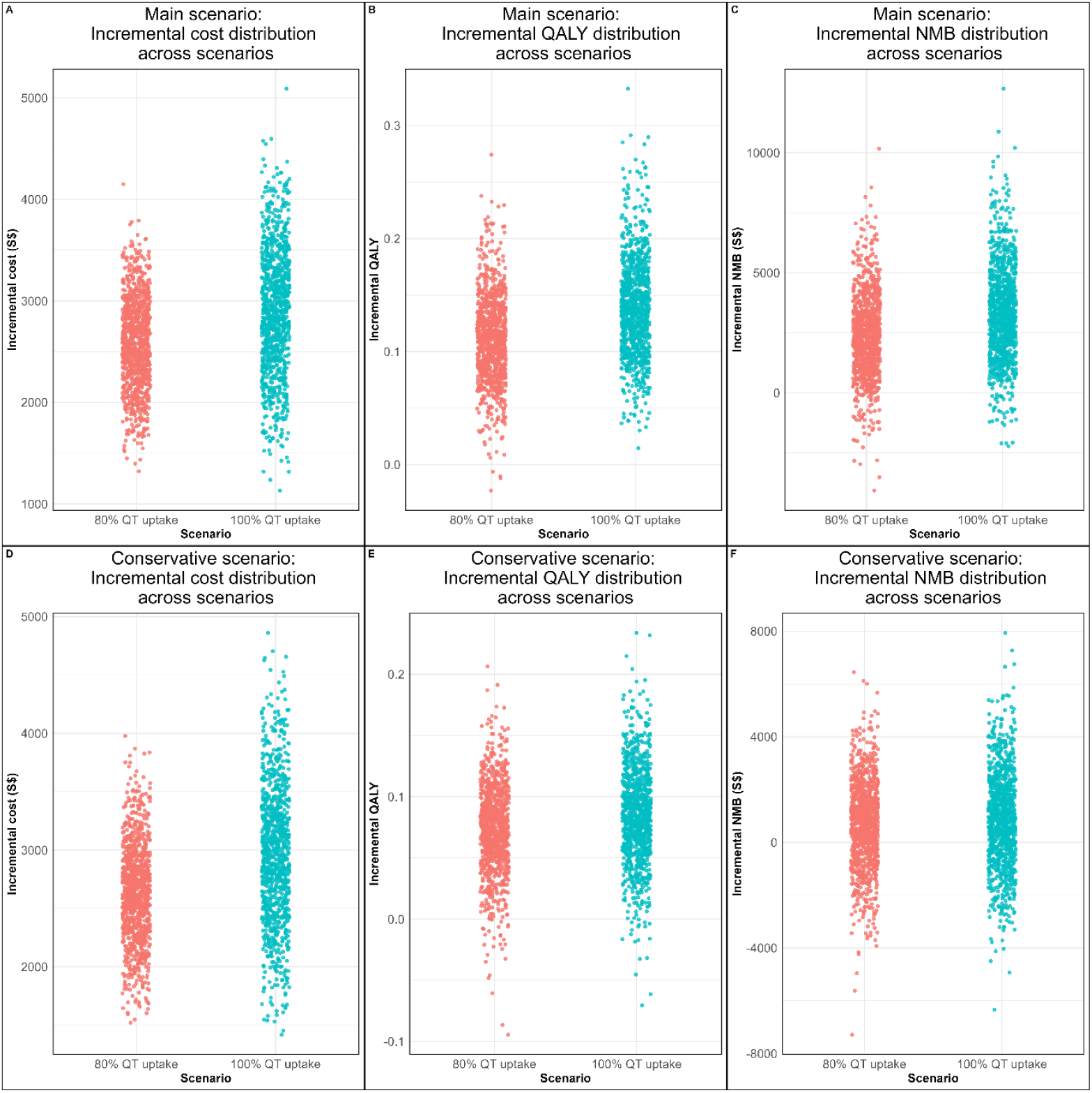
Incremental cost, incremental effectiveness and incremental net monetary benefit scatterplots for the realistic (80%) and stretch (100%) quadruple-therapy scale-up scenarios, showing results from 1,000 Monte Carlo simulations

## Discussion

This model-based analysis demonstrates that increased use of quadruple therapy for HFrEF in Singapore is a highly cost-effective strategy. Under both the realistic (80% uptake) and stretch (100% uptake) scale-up scenarios, the incremental cost per QALY gained remains well below Singapore’s willingness-to-pay threshold ($45,000/QALY). Even when we applied conservative assumptions, quadruple therapy scale-up continued to be cost-effective. These findings underscore that investing in comprehensive GDMT delivery for HFrEF delivers substantial health gains with good value for money.

Notably, our findings align with prior economic evaluations from other healthcare settings. One U.S. study estimated that comprehensive quadruple therapy yields approximately 1.1 additional QALYs per patient compared to conventional therapy (ACEi, beta-blocker, and MRA), with an ICER of around US$81,000 per QALY—well within acceptable U.S. value thresholds^10^. Another recent U.S. analysis assessed the sequential addition of SGLT2i and ARNi to standard therapy and found that full quadruple therapy generated 0.68 additional QALYs over SGLT2i alone, with an ICER of US$98,500 per QALY^11^. These studies collectively underscore that optimising HFrEF management with all four drug classes substantially improves survival and quality of life, offering good economic value and supporting broader implementation efforts. Our Singapore-specific findings echo this global evidence: increased adherence to quadruple therapy is a high-value intervention that warrants policy attention.

Translating these favourable cost-effectiveness findings into practice will require addressing real-world implementation barriers within Singapore’s healthcare system. At present, only around 30% of an eligible patient cohort with HFrEF was receiving quadruple therapy, leaving considerable scope for improvement. Several factors likely contribute to this gap and should be further evaluated. Clinician inertia and limited familiarity with newer therapies may delay uptake, particularly where competing priorities exist in busy clinical settings. Patients with HFrEF often have multiple comorbidities and are already on complex medication regimens, making the initiation and titration of four evidence-based drug classes logistically challenging. This raises concerns around polypharmacy, adherence, and the need for close monitoring. Cost and access considerations may also influence uptake. While Singapore’s public healthcare system offers drug subsidies, newer HFrEF therapies may still incur higher out-of-pocket costs, potentially deterring their use. Ensuring affordability at the point of care will be important to avoid financial barriers. Another critical issue is system capacity to support appropriate titration. Quadruple therapy often requires regular and more frequent follow-up for monitoring blood pressure, renal function, and electrolytes, as well as patient education to manage side effects. Embedding these activities into routine care may require additional resources, for example, strengthening pharmacist and nurse-led titration clinics that have been successfully implemented in other countries^19,20^. While increasing adherence from 30% to 80% uptake is ambitious, it may be achievable with targeted, system-level interventions that address clinical, organisational, and financial barriers.

Singapore can draw important insights from international experiences to improve the uptake of GDMT for HFrEF. In the United States, the American Heart Association’s IMPLEMENT-HF initiative demonstrated that large-scale improvements are achievable^21^. Through collaborative learning, real-time audit-and-feedback, and attention to social needs, participating hospitals increased quadruple therapy use at discharge nearly ten-fold from 4.7% to 44.6%. Within 30 days post-discharge, uptake rose to about 45%, showing that system-level interventions can overcome low baseline usage. Routine assessment of patients for potential social determinants of health that are barriers to accessing GDMT and health resources can be considered.

Timing of therapy initiation also matters. Starting GDMT during hospitalisation has been linked to significantly higher long-term adherence; in contrast, delayed initiation often results in patients remaining untreated for extended periods^22^. This highlights the importance of embedding medication initiation into hospital discharge protocols, supported by early follow-up. Furthermore, multidisciplinary care models, such as team-based heart failure clinics involving nurses, pharmacists, and cardiologists have shown success in optimising therapy and reducing readmissions. Nurse-led titration clinics, in particular, enable safe and efficient medication adjustments. Allied health led clinics and teleconsultation systems are in place in some Singapore institutions and could be up-scaled to support the timely and equitable titration of quadruple therapy.

In our analysis we assumed a one-time intervention cost of $1,000 per patient to achieve 80% uptake of quadruple therapy. The exact cost and effectiveness of such an intervention are currently unknown. However, our results suggest that any strategy capable of achieving this compliance level at or below this cost would be a cost-effective option. This can inform the design of future implementation trials structured around predefined effectiveness and cost thresholds that directly reflect decision-makers’ needs. Such a design would align evidence generation with real-world resource constraints and enable more agile and policy-relevant evaluation.

This study has several limitations. First, due to limited patient numbers and follow-up time in the local dataset, we were unable to estimate transition probabilities for the quadruple therapy cohort directly from Singapore-specific data. Instead, treatment effects were derived from published international trials, which may not fully reflect the local population or healthcare context. Second, while we incorporated local data for hospitalisation costs and drug regimens, some key parameters such as utility values were sourced from regional or global literature due to the absence of Singapore-specific estimates. This may affect the generalisability of our findings. Third, our model assumed patients remained on a fixed drug regimen throughout the 10-year horizon without accounting for potential treatment discontinuation, switching, or titration, which are common in clinical practice and could influence real-world cost and effectiveness outcomes. Finally, behavioural and adherence-related factors were not explicitly modelled. In practice, medication adherence, patient education, and clinician prescribing behaviour significantly influence treatment effectiveness, and their omission may lead to overestimation of the achievable benefits of quadruple therapy in routine care.

In conclusion, increasing adherence to quadruple therapy for HFrEF in Singapore is a clinically impactful and a potentially cost-effective policy decision. Both the 80% and 100% uptake scenarios demonstrated the criterion for cost-effectiveness was met, reinforcing the need to close the current treatment gap. Achieving this will require coordinated policy, system-level support, and investment in implementation strategies. With the right actions, Singapore can substantially improve heart failure outcomes, reduce hospitalisations, and deliver greater value to patients and the healthcare system alike.

## Data Availability

The data used in this study were obtained from the Singapore Cardiovascular Longitudinal Outcomes Database (SingCLOUD). Access to these data is restricted, and they are not publicly available. Interested researchers can obtain access to the data by contacting the managing team of SingCLOUD for further information.

## Contributors

*Study concept and design:* Sameera Senanayake, Audry Shan Yin Lee, Sanjeewa Kularatna, Nicholas Graves

*Data curation:* Annie Lee, Yee How Lau

*Funding acquisition:* Derek J Hausenloy, Mark Yan-Yee Chan

*Data analysis:* Sameera Senanayake

*Data interpretation:* Sameera Senanayake, Audry Shan Yin Lee, Annie Lee, Yee How Lau, Derek J Hausenloy, Khung-Keong Yeo, Mark Yan-Yee Chan, Raymond Ching Chiew Wong, Seet Yoong Loh, Kheng Leng David Sim, Phoo Pyae Sone Win, Kelvin Bryan Tan

*Writing—original draft:* Sameera Senanayake

*Writing—review & editing:* Nicholas Graves, Audry Shan Yin Lee, Annie Lee, Derek J Hausenloy, Khung-Keong Yeo, Mark Yan-Yee Chan, Raymond Ching Chiew Wong, Seet Yoong Loh, Kheng Leng David Sim, Chow Weien, Thin Mar Win, Kelvin Bryan Tan

*Supervision:* Nicholas Graves

All authors read and approved of the final manuscript. All authors accept responsibility for submitting the final manuscript for publication.

## Acknowledgements

DJH is supported by the Duke-NUS Signature Research Programme funded by the Ministry of Health, Singapore Ministry of Health’s National Medical Research Council under its Singapore Translational Research Investigator Award (MOH-STaR21jun-0003), Centre Grant scheme (NMRC CG21APR1006), and Collaborative Centre Grant scheme (NMRC/CG21APRC006). This article is based on work supported by the CVD Health Services Research (HSR) Unit under the CArdiovascular DiseasE National Collaborative Enterprise (CADENCE) National Clinical Translational Program (MOH-001277-01). EditGPT was used for language editing and proofreading purposes during manuscript preparation. No content, analysis, or interpretation was generated by AI. All substantive content and intellectual input were provided by the authors.

## Reference

1. Akinterinwa O, Singh M, Vemuri S, Tyagi S. A Need to Preserve Ejection Fraction during Heart Failure. International Journal of Molecular Sciences. 2024-08-01 2024;25doi:10.3390/ijms25168780

2. Reddy Y, Borlaug B. Heart Failure With Preserved Ejection Fraction. Current problems in cardiology. 2016-04-01 2016;41 4:145–188. doi:10.1016/j.cpcardiol.2015.12.002

3. Senanayake S, Kularatna S, Lee ASY, et al. Health Services Costs of Clinical Heart Failure With Reduced Ejection Fraction in Singapore. Value in Health Regional Issues. 2025;45:101037.

4. Lam C, Gamble G, Ling L, et al. Mortality associated with heart failure with preserved vs. reduced ejection fraction in a prospective international multi-ethnic cohort study. European Heart Journal. 2018-05-21 2018;39:1770. doi:10.1093/eurheartj/ehy005

5. McMurray JJ, Packer M, Desai AS, et al. Angiotensin–neprilysin inhibition versus enalapril in heart failure. New England Journal of Medicine. 2014;371(11):993–1004.

6. McMurray JJ, Solomon SD, Inzucchi SE, et al. Dapagliflozin in patients with heart failure and reduced ejection fraction. New England Journal of Medicine. 2019;381(21):1995–2008.

7. Tang A, Ziaeian B, Butler J, Yancy C, Fonarow G. Global Impact of Optimal Implementation of Guideline-Directed Medical Therapy in Heart Failure. JAMA cardiology. 2024-10-02 2024;doi:10.1001/jamacardio.2024.3023

8. Tan CC, Lam CS, Matchar DB, Zee YK, Wong JE. Singapore’s health-care system: key features, challenges, and shifts. The Lancet. 2021;398(10305):1091–1104.

9. McDonagh TA, Metra M, Adamo M, et al. 2021 ESC Guidelines for the diagnosis and treatment of acute and chronic heart failure: Developed by the Task Force for the diagnosis and treatment of acute and chronic heart failure of the European Society of Cardiology (ESC) With the special contribution of the Heart Failure Association (HFA) of the ESC. European heart journal. 2021;42(36):3599–3726.

10. Dixit NM, Parikh NU, Ziaeian B, Jackson N, Fonarow GC. Cost-effectiveness of comprehensive quadruple therapy for heart failure with reduced ejection fraction. Heart Failure. 2023;11(5):541–551.

11. Yan BW, Spahillari A, Pandya A. Cost-effectiveness of quadruple therapy in management of heart failure with reduced ejection fraction in the United States. Circulation: Cardiovascular Quality and Outcomes. 2023;16(6):e009793.

12. Yeo KK, Ong H-Y, Chua T, et al. Building a longitudinal national integrated cardiovascular database-lessons learnt from SingCLOUD-. Circulation Reports. 2020;2(1):33–43.

13. Zhang Z. Parametric regression model for survival data: Weibull regression model as an example. Annals of translational medicine. 2016;4(24):484.

14. Allison PD. Survival analysis using SAS: a practical guide. Sas Institute; 2010.

15. Vaduganathan M, Claggett BL, Jhund PS, et al. Estimating lifetime benefits of comprehensive disease-modifying pharmacological therapies in patients with heart failure with reduced ejection fraction: a comparative analysis of three randomised controlled trials. The Lancet. 2020;396(10244):121–128.

16. Kularatna S, Byrnes J, Chan YK, Carrington MJ, Stewart S, Scuffham PA. Comparison of contemporaneous responses for EQ-5D-3L and Minnesota Living with Heart Failure; a case for disease specific multiattribute utility instrument in cardiovascular conditions. International journal of cardiology. 2017;227:172–176.

17. McMurray JJ, Trueman D, Hancock E, et al. Cost-effectiveness of sacubitril/valsartan in the treatment of heart failure with reduced ejection fraction. Heart. 2018;104(12):1006–1013.

18. See-Toh RS-E, Wong XY, Mahboobani KSKH, et al. Cost-effectiveness of transcatheter aortic valve implantation in patients with severe symptomatic aortic stenosis of intermediate surgical risk in Singapore. BMC health services research. 2022;22(1):994.

19. Freedman G, Watt R, Chowdhury EK, et al. Nurse-Led, Remote Optimisation of Guideline-Directed Medical Therapy in Patients with Heart Failure and Reduced Ejection Fraction Across Australia. Journal of Clinical Medicine. 2025;14(15):5371.

20. Zheng J, Mednick T, Heidenreich PA, Sandhu AT. Pharmacist-and nurse-led medical optimization in heart failure: a systematic review and meta-analysis. Journal of cardiac failure. 2023;29(7):1000–1013.

21. Sauer AJ, Beon C, Cherkur S, et al. Multiregional Implementation Initiative’s Impact on Guideline-Based Performance Measures for Patients Hospitalized With Heart Failure: IMPLEMENT-HF. Circulation: Heart Failure. 2025;18(5):e012547.

22. Mebazaa A, Davison B, Chioncel O, et al. Safety, tolerability and efficacy of up-titration of guideline-directed medical therapies for acute heart failure (STRONG-HF): a multinational, open-label, randomised, trial. The Lancet. 2022;400(10367):1938–1952.

